## Supplementary Materials for "COVID-19: Short term prediction model using daily incidence data"

### Appendix A. Methods

To model the number of cases, we use the simplest time-since-infection model which is also known as the "Kermack-McKendrick" model [1]. Following similar setup from Fraser [2] and Cori et al. [3], we define a function $\beta(t,\tau)$ which is the transmission probability at a calendar time $t$, after being infected $\tau$ time ago. Therefore, between time $t$ and $t+\delta$, someone infected at time $\tau$ ago will consequently infect someone else with a probability of $\beta(t,\tau)\delta$. As a result, the number of cases at time $t$, $I(t)$, is related back to previous cases through the following renewal equation:

$$I(t) = \int_0^t \beta(t,\tau) I(t-\tau) \mathbf{d}\tau. \tag{1}$$

Suppose that $\beta(t,\tau)$ is separable, that is:

$$\beta(t,\tau) = R_e(t) w(\tau),$$

where, with no loss of generality, we assume

$$\int_0^\infty w(\tau)\mathbf{d}\tau = 1,$$

such that

$$R_e(t) = \int_0^\infty \beta(t,\tau)\mathbf{d}\tau.$$

This function $R_e(t)$ therefore estimates the number of people someone infected at time $t$ would infect if the conditions of the epidemic remains constant. It was called "instantaneous reproduction number" by Cori et al. [3]. In this paper, we refer to this parameter as the effective reproduction number in order to distinguish it from the basic reproduction number $R_0$ – the average number of people infected by one infectious person under a natural situation when no intervention is implemented. The effective reproduction number $R_e$ is a very important parameter, since it can be shown that when $R_e$ is greater than one, infections will increase and lead to an outbreak where more and more people will be infected. If $R_e$ is less than one, then the outbreak will die down eventually. The effective reproduction number can be influenced by four major factors [4]: 1) transmissibility of the disease; 2) duration of the infectious period; 3) number of contacts between infectious and susceptible individuals each day; and 4) the percentage of people that are immune or no longer susceptible. We will show that this parameter plays a major role in our forecasting model for COVID-19.

The normalized function $w(\tau)$ represents the relative transmissivity at a time $\tau$ after infection. The distribution function $w(\tau)$, also called the distribution of the serial interval, can be obtained only if the infection date for each case is known. In a real world application, we usually only have the information of the reported dates of each case, instead of the infection date, therefore, we will rely on either the external source of data or we will make some assumptions about the serial distribution $w(\tau)$.

Plugging in the factorization for $\beta(t,\tau)$, the renewal function (1) becomes

$$I(t) = R_e(t) \int_0^t I(t-\tau) w(\tau)\mathbf{d}\tau.$$

In practice, incidence data are obtained usually at daily intervals, and thus are discrete. Therefore, the discrete version of the above renewal equation is

$$I(t) = R_e(t) \sum_{j=1}^{t} I(t-j) w(j), \qquad t = 1, 2, \cdots, T, \tag{2}$$

where $T$ indicates the last time point of the incidence series.

Even though we could obtain an estimate of $R_e$ from the above equation, the estimate can be highly variable, due to reporting errors (for example, under-reporting and delayed reporting) associated with the incidence case series. Therefore, we follow the setup of Cori et al. [3] and view the cases as arising from a Poisson Process, where the mean parameter is given by the renewal equation (2). In order to obtain a more stable estimate of $R_e(t)$, we also assume that $R_e(t)$ is the same for a total of $\tau$ days during the period $[t-\tau+1, t]$. Explicitly, the likelihood function of the cases during the time period $[t-\tau+1, t]$ is as follows:

$$Pr(I_{t-\tau+1}, \cdots, I_t | I_0, I_1, ..., I_{t-\tau}, w, R_e(t)) = \prod_{j=t-\tau+1}^{t} \frac{e^{-R_e(t)\Lambda_j} \{R_e(t)\Lambda_j\}^{I_j}}{I_j!},$$

where

$$\Lambda_j = \sum_{s=1}^{j} I_{j-s} w(s).$$

Following Cori et al. [3], we will use a Bayesian framework and assume $R_e(t)$ has a prior Gamma distribution with shape parameter $a$ and scale parameter $b$. Since we have a Poisson likelihood, a gamma prior is conjugate, and should result in a gamma posterior for $R_e(t)$ with shape parameter $a^*$ and scale parameter $b^*$, where

$$a^* = a + \sum_{j=t-\tau+1}^{t} I_j, \quad b^* = \frac{1}{\frac{1}{b} + \sum_{j=t-\tau+1}^{t} \Lambda_j}. \tag{3}$$

For now, we set $a$ and $b$ to be hyper parameters specified by the user. We defer future discussion of these parameters and the serial interval function $w(s)$ to the next section.

This procedure can now be used to get the posterior distribution of $R_e(t)$ for any time interval $[t-\tau+1, t]$. The interval length $\tau$ is chosen such that there is a large enough data set to provide stable estimates, and small enough to capture the time-varying nature of $R_e(t)$. Eventually, we will use the posterior distribution of $R_e(T)$ to predict future cases, where $T$ is the last time point of the observed incidence series.

For short-term forecasting of new cases, we propose a method that uses very few assumptions and is therefore straightforward to model. We simply assume that in the near future after time $T$, $R_e(t)$ will 1) stay the same; 2) increase 5%; 3) decrease 5%. For the first scenario that $R_e(t)$ will stay the same, we perform the following procedures for making predictions:

1. Draw a new $R_e^*$ from the posterior gamma distribution of $R_e(T)$, as specified by (3), where $T$ is the end time point of the observed incidence series.

2. Then we draw $I_{T+1}$ from a Poisson distributing with a mean function obtained by the renewal function (2).

3. Similarly, we draw sequentially the values for $I_{T+2}, I_{T+3}, \cdots, I_{T+K}$, where $K$ is predefined forecasting period. Thus we obtain one complete forecast series, $I_{T+1}, I_{T+2}, \cdots, I_{T+K}$.

4. Repeat steps (1) through (3), and obtain multiple sample series, from which we can get predicted means $I_t^*$, and the predicted 95% confidence intervals $[I_t^{L*}, I_t^{U*}]$, $t = T+1, \cdots, T+K$.

Since the reproduction number $R_e$ may change with policy implementation and public behavior, we also predict the scenarios when $R_e$ changes by $\delta$, for example,

$\delta = 5\%$, or $\delta = -5\%$. Either way, we borrow the posterior distribution obtained from $R_e(T)$, and assume that the mean parameter changes by $\delta$, but the variance stays the same. This is equivalent to setting the posterior distribution for $R_e$ to be a gamma distribution with shape parameter $a^*_\delta$ and scale parameter $b^*_\delta$, where $a^*_\delta = a^*\delta^2$, $b^*_\delta = b^*/\delta$. Then we follow the same steps outlined above to predict the means and the 95% confidence intervals for $I_{T+1}, I_{T+2}, \cdots, I_{T+K}$.

#### Appendix B. Choosing Model Parameters

The goal of our proposed method is to make as few approximations as possible, so that we can make reasonable predictions without having to rely on the correct specification of the assumptions. However, given only the incidence data, the reproduction number $R_e(t)$ can not be determined uniquely [4], so some basic assumptions are necessary. The most important assumption is the serial interval distribution. Based on literature reviews, we adopted a discretized gamma distribution [3]. The mean of 3·95 days and a standard deviation of 4·24 days were based on the work of Ganyani et al. [5] using COVID-19 data from Tianjin, China. If desired, a prior can be placed on the parameters for the mean and standard deviation of the serial interval, and this approach was discussed in estimating $R_e(t)$ values in Cori et al. [3].

There are other hyper parameters that must be supplied. In our method, we assume that each $R_e(t)$ has a gamma prior distribution, with shape and scale parameters $a$ and $b$. It is common knowledge that the posterior distribution of $R_e(t)$ depends both on the parameters in the prior distribution, and the observed data. When the incidence number is relatively high, as experienced in Texas and other states during the Summer and Fall of 2020, the prior parameters have very little effect on the final posterior distribution. Our experience suggests that a wide range of choices worked for $a$ and $b$ as hyper-parameters. Choosing a single $a$ and $b$ and using the same prior in general for all $R_e(t)$ work well. We have chosen a value of $a = 1, b = 5$ following the work of Cori et al. [3]. Another strategy is to choose the prior parameters so that the prior has a mean that is equal to the previous estimate $R_e(t - \tau)$, and a standard deviation resembling the posterior standard deviation of the previous $R_e(t - \tau)$.

The last parameter we need to decide is the interval parameter $\tau$, since we based our prediction on the estimated $R_e(T)$ during the interval $[T - \tau + 1, T]$, where $T$ is the end time of a time series upon which we wish to make a forecast. The choice of $\tau$ is a compromise between the variability and the accuracy of the predictions. In general, choosing $\tau$ to be small will result in highly variable estimates of $R_e(t)$, but will be more accurate due to the fact that $R_e(t)$ may be different at different times. In contrast, a large $\tau$ is less accurate, but also less variable, as it uses more data to fit $R_e(t)$. In our experience, a choice between 7 and 12 days worked well when we applied them to the real data sets.

We end this section on a note of using a Poisson distribution for the likelihood of the observed incidence sequence. Despite the over-dispersion issue associated with the Poisson distribution assumptions (i.e. the Poisson distribution assumes that the variance is the same as the mean, but in reality, the variance is often larger than the mean), our experience is that the predicted cases can vary a lot due to the change of the underlying reproduction number $R_e(t)$. By allowing $R_e(t)$ to take different values in the prediction, we will capture the uncertainty of the future incidence series in a different way. Our prediction interval will be determined by different transmission rate scenarios: the upper bound is given by the upper 95% confidence limit when the transmission rate increase 5%, and the lower bound is given by the lower 95% confidence interval limit when the transmission rate decreases 5%.

#### References


1. Kermack W, McKendrick A. A contribution to the Mathematical Theory of Epidemics. *Proc Royal Soc London A*. 1927;**115**:700–721.

2. Fraser C. Estimating individual and household reproduction numbers in an emerging epidemic. *PloS one*. 2007;**2**(8):e758.

3. Cori A, Ferguson NM, Fraser C, Cauchemez S. A new framework and software to estimate time-varying reproduction numbers during epidemics. *American journal of epidemiology*. 2013;**178**(9):1505–1512.

4. Britton T, Scalia Tomba G. Estimation in emerging epidemics: Biases and remedies. *J R Soc Interface*. 2019;**16**:20180670.

5. Ganyani T, Kremer C, Chen D, Torneri A, Faes C, Wallinga J, et al. Estimating the generation interval for coronavirus disease (COVID-19) based on symptom onset data, March 2020. *Eurosurveillance*. 2020;**25**(17):2000257.
